# A significant role for maternal genetic nurture in the risk architecture of attention-deficit/hyperactivity disorder

**DOI:** 10.1101/2024.12.23.24319549

**Authors:** Behrang Mahjani, Adrianna P. Kępińska, Shelby Smout, Lily Cohen, Madison Caballero, Joseph D. Buxbaum, Dorothy E. Grice

**Affiliations:** Seaver Autism Center for Research and Treatment, Icahn School of Medicine at Mount Sinai, New York, NY, USA; Department of Psychiatry, Icahn School of Medicine at Mount Sinai, New York, NY, USA; Department of Genetics and Genomic Sciences, Icahn School of Medicine at Mount Sinai, New York, NY, USA; Department of Artificial Intelligence and Human Health, Icahn School of Medicine at Mount Sinai, New York, NY, USA; The Mindich Child Health and Development Institute, Icahn School of Medicine at Mount Sinai, New York, NY, USA; Department of Molecular Medicine and Surgery, Karolinska Institutet, Stockholm, Sweden; Department of Medical Epidemiology and Biostatistics, Karolinska Institutet, Stockholm, Sweden; Friedman Brain Institute, Icahn School of Medicine at Mount Sinai, New York, NY, USA; Department of Neuroscience, Icahn School of Medicine at Mount Sinai, New York, NY, USA; Division of Tics, OCD and Related Disorders, Icahn School of Medicine at Mount Sinai, New York, NY, USA

## Abstract

**Objective:** Previous studies have shown that parental factors are associated with an increased risk for attention-deficit/hyperactivity disorder (ADHD). However, the pathways by which parental factors are associated with risk of ADHD in offspring are not well understood. These associations can arise directly from parental genotypes inherited by offspring, and/or via environmental effects, some of which may themselves be influenced by the parental genotype (i.e., parental genetic nurture). This study specifically examines the impact of the maternal phenotype on offspring ADHD risk, above and beyond the direct genetic effect of maternally-inherited genes.

**Methods:** The study population consisted of 982,544 individuals from the Swedish Medical Birth Register. We partitioned the liability of ADHD into direct additive genetic effect, maternal genetic nurture effect, and maternal common environment effect.

**Results:** We identified 66,707 (7%) individuals in the birth cohort with an ADHD diagnosis. Maternal half-siblings were associated with a higher ADHD risk compared to paternal half-siblings, suggesting maternal effect. We estimated 66.1% direct additive genetic effect (95% credible interval, 0.647%-0.676%) and 14.3% maternal genetic nurture effect (95% credible interval, 0.136%-0.151%). Additionally, we also observed evidence for substantial assortative mating among individuals with ADHD.

**Conclusions:** Our findings indicate that maternal genotypes influence offspring’s risk of ADHD through environmental pathways beyond the effects of direct genetic transmission. Exploring the impact of the genetics of the mother beyond the maternally inherited genes can lead to new insights into ADHD risk. Future studies should also investigate paternal effects to provide a more comprehensive understanding of the ADHD risk architecture.

## 1. Introduction

Attention-deficit/hyperactivity disorder (ADHD) is a neuropsychiatric disorder characterized by persistent patterns of inattention and/or hyperactivity and impulsivity that interfere with an individual’s daily functioning and neurodevelopment (1). The prevalence of ADHD is 7.6% for children aged 3 to 12 years (95% confidence interval: 6.1–9.4%) and 5.6% for teenagers aged 12 to 18 years (95% confidence interval: 4.8-7%) (2).

ADHD is thought to arise from a complex interplay of genetic and environmental risk factors (3, 4). Evidence from twin, family, and adoption studies shows that ADHD is highly heritable, with estimates ranging from 50% to 80% (5–8), indicating that genetic factors play a significant role in its development (9). Environmental risk factors for ADHD have been extensively evaluated (10–12), with research proposing that the maternal environment, before, during, and post-pregnancy, may significantly affect the risk of developing ADHD in offspring (13–15). For example, maternal pre- and perinatal exposure to toxins and pollutants, diet, psychosocial stressors, smoking, and alcohol consumption are all possible risk factors (16–21). Studies have also shown that a variety of perinatal maternal psychiatric disorders are positively associated with ADHD, such as maternal stress and anxiety, maternal prenatal stress, maternal depression, and maternal post-partum depression (22, 23).

The association between maternal factors and the risk of ADHD could be explained by confounding environmental factors and/or the maternal genotype inherited by the offspring (genetic confounding) (3). We posit these maternal factors could increase the risk of ADHD in offspring via three different pathways, as illustrated by arrows 1, 2, and 3 in Figure 1. The first pathway (arrow 1) involves the direct transmission of genes (direct genetic effect) from the maternal genome to the offspring genome. The second and third pathways (arrows 2 and 3), known as *maternal effect*, are the influence of maternal phenotype on the phenotype of the offspring above and beyond the transmission of maternal genes to the offspring, i.e., direct genetic inheritance (24). Maternal effect can arise from the maternal genotype (maternal genetic nurture effect, also referred to as genetic maternal effect, arrow 2), the maternal environment (maternal common environment effect, also known as environmental maternal effect, arrow 3), or their combined effects.

**Figure 1.**
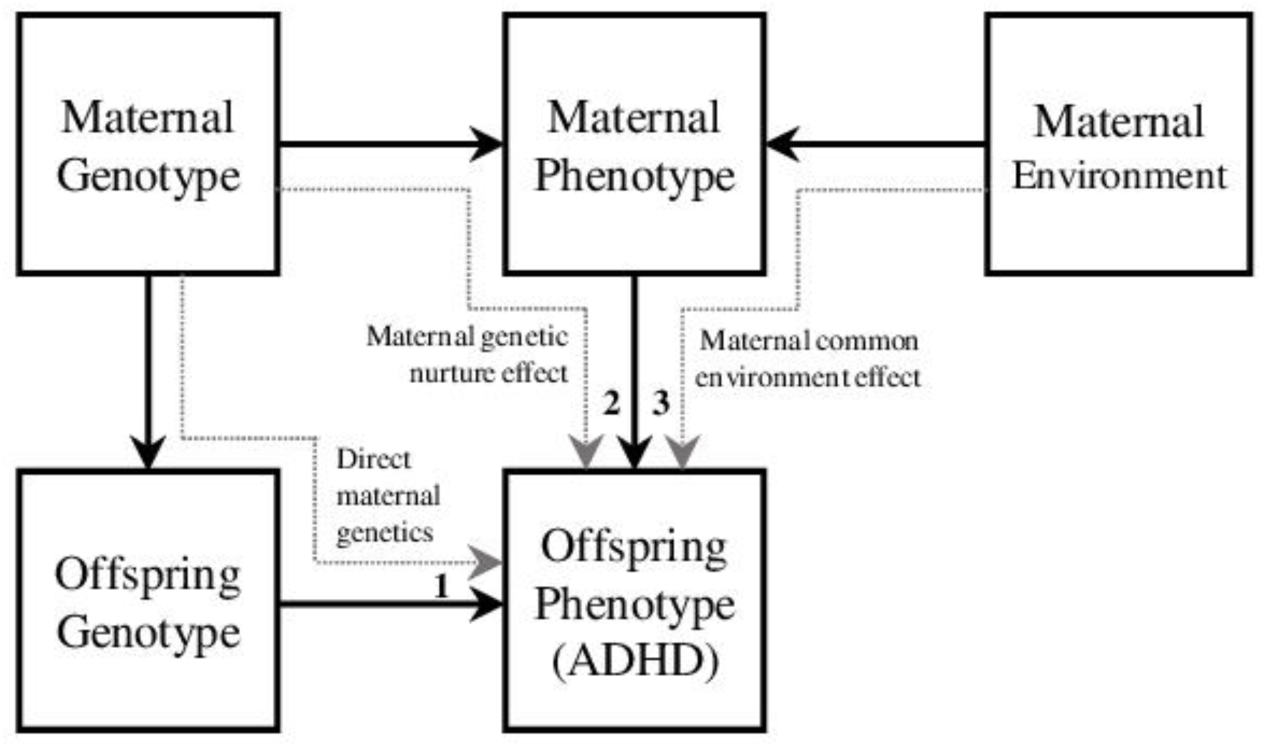
An illustration demonstrating maternal effect. We demonstrate the maternal genetic nurture effect between mother and offspring (maternal genotype to maternal phenotype, and then maternal phenotype to offspring phenotype, dashed arrow 1), the maternal common envrionment effect (maternal environment to maternal phenotype, and then maternal phenotype to offspring phenotype, dashed arrow 2), and direct maternal genetic effect (maternal genotype to offspring genotype, offspring genotype to offspring phenotype, dashed arrow 3). Paternal effect, child environment effect and gene×environment interactions are not illustrated in this figure.

Maternal effect falls under the category of indirect genetic effects, which refer to genetic effects on a trait that are not transmitted through direct genetic inheritance but rather through the influence of an individual’s genotype on the phenotype of another individual (25). Other forms of indirect genetic effects, such as horizontal indirect genetic effects (i.e., within-generation indirect genetic effects)(26), have been observed in studies of behavioral phenotypes, such as educational attainment among school friends (27).

Paternal effect could also contribute to the offspring’s risk of ADHD. Although fathers do not directly influence the fetal prenatal environment as mothers do, paternal factors can still impact offspring health through their effects on the maternal or child environment and behavior. For example, paternal smoking or stress levels could potentially influence maternal stress and health during pregnancy, indirectly affecting the fetus. However, incorporating paternal effects into the model significantly complicates the analysis due to the need to account for interactions between maternal and paternal influences, which may have non-additive effects on the offspring’s phenotype.

While many studies have determined different maternal risk factors for ADHD (3, 4, 28, 29), a knowledge gap remains between how these factors arise and their role on the phenotype of the offspring. Our preliminary observations suggest that relatives connected through maternal lines— such as maternal half-siblings and maternal cousins—tend to be more similar in their ADHD risk than those related through paternal lines. This pattern motivated our analyses, implying that maternal factors, whether genetic, environmental, or both, may uniquely shape the familial transmission of ADHD. Here, we address to fill this gap by estimating the role of maternal effect on risk of ADHD in offspring using a large population-based prospectively ascertained cohort of Swedish-born individuals. We assessed the robustness of estimates of direct additive genetics and maternal effects under different models, as described in the supplementary material.

Assortative mating has been reported among individuals with a diagnosis of ADHD (30). Assortative mating can increase the estimate of population prevalence and inflate the estimate of similarities between relatives. Therefore, we also evaluated the influence of assortative mating on the estimation of direct additive genetics.

## 2. Material and methods

### 2.1 Study population

All data were sourced from the Swedish national registers. Sweden has universal health care coverage that is publicly financed and available to all citizens. Since 1973, all children born in Sweden have been recorded in the National Medical Birth Register, together with the birth characteristics of the children and mothers. Quality controls of registers are performed at a national level on the submitted data and include checking the quality and validity of the entries, such as personal registration number, hospital, and diagnoses. Data suspected of containing erroneous or invalid data points is followed up and corrected.

Our source population included all liveborn singleton children born in Sweden between January 1, 1992, and December 31, 2002, whose mother and father were recorded in the National Medical Birth Register. Prospective follow-up continued until December 2018. We excluded individuals who emigrated during the study period to ensure complete information on the diagnosis. The data from the Swedish Multi-Generation Register were used to establish family relationships by providing details about each child’s relatives.

Ethical approval and waiver of informed consent were obtained from the Regional Ethical Review Board in Stockholm.

### 2.2 Outcomes and Exposure Covariate

All psychiatric hospital admissions in Sweden occurring after 1973 are recorded in the National Patient Register. Beginning in 2001, outpatient specialist care was also recorded. Diagnoses in the Swedish National Patient Register were made by a specialist, in this case, by a psychiatrist at a hospital. After a diagnosis was made, the diagnoses were registered using the International Classification of Diseases (ICD) codes.

Diagnostic information about ADHD was obtained using data from the National Patient Register, based on ICD-10: F90. We used sex at birth as a fixed effect (covariate) in the analyses.

### 2.3 Statistical Analysis

We used a *family-based quasi-experimental design* that leverages on ‘natural experiments’ arising from inherent differences in family relationships (31, 32). These differences include variations in genetic relatedness, such as between full siblings, half-siblings, and cousins, and differences in environmental exposure within the same household. This method allows us to observe the effects of genetic and environmental factors in a quasi-controlled setting, closely resembling the conditions of randomized controlled trials. By exploiting these naturally occurring variations among family members, we can more effectively control for confounding variables and enhance the validity of our causal estimates. Family-based designs are particularly effective in controlling for genetic confounding and improving causal inference in observational.

We defined family relationships based on first, second, and third-degree relatives: full siblings (FS), paternal and maternal half-siblings (pHS, mHS), and three different cousin types, depending on whether the two parents responsible for the cousin relationship are 1) sisters: maternal parallel cousins (mPC), 2) brothers: paternal parallel cousins (pPC), or 3) brother and sister: cross cousins (CC). Contrasts of recurrence risk for mHS versus pHS and mPC versus pPC and CC can be utilized to estimate maternal effect because additive genetic contributions are predictable, and shared environmental effects should be minimized and are assumed to be zero in our models for cousins.

We used all possible pairs of siblings and cousins with available family information, born between January 1, 1992, and December 31, 2002. We have previously used this approach to study the role of maternal effect on the risk of obsessive-compulsive disorder (OCD) (33) and Tourette syndrome (34).

We estimated the relative recurrence risk of ADHD for different relationship types using Cox proportional hazards regression, with attained age as the primary time scale and adjusted for sex (assigned at birth). Each individual was followed from birth until either death, emigration from Sweden, an ADHD diagnosis, or the end of follow-up on December 31, 2018.

The estimates of relative recurrence risks derived from different family types were utilized to estimate direct additive genetics and maternal effect on ADHD (supplementary material, section S1). Table 1 provides a detailed summary of how phenotypic covariance contributions can be partitioned across direct genetic (DG), maternal genetic nurture effect (MGN), and maternal common environment effect (MCE) for each type of relationship considered in this study. These parameters can be estimated using the Falconer threshold liability method (35), which involves comparing the tetrachoric correlation between siblings and cousins with a random sample obtained from the general population. The Falconer method has been one of the primary approaches for estimating heritability (see supplementary material, section S1). However, this method cannot account for data with different family sizes (unbalanced sampling designs). Generalized linear mixed models (GLMM) using restricted maximum likelihood have been a major change in the study of quantitative genetics (chapter 28, (36)) because GLMMs can account for varying family sizes and unbalanced samples (37). Ideally, the results obtained from the family-matched design and the GLMM should converge (38).

**Table 1.** Partitioning of phenotypic covariance contributions among relatives by direct genetic effect (DG), maternal genetic nurture effect (MGN), and maternal common envrionment effect (MCE)

| Relationship type | DG | MGN | MCE | Explanation |
| --- | --- | --- | --- | --- |
| **Full Siblings (FS)** | 0.5 | 1 | 1 | ▪ The coefficient for DG represents the coefficient of genetic relationship, which is twice the probability that two gametes sampled at random from each parent will carry alleles identical by descent (come from the same ancestor). Full siblings share, on average, 50% of their genes, leading to a DG of 0.5.<br>▪ The coefficient for MGN represents the coefficient of genetic relationship through maternal linkage. For full siblings, MGN is set to 1 because they share the same biological mother.<br>▪ The coefficient for MCE represents the coefficient of the shared environment provided by the mother. For full siblings, MCE is set to 1 because they share the same biological mother. |
| **Maternal Half-Siblings (mHS)** | 0.25 | 1 | 1 | ▪ For half-siblings, the coefficient for DG is half of the DG for full siblings because they only share one biological parent.<br>▪ MGN is set to 1 because mHS share the same biological mother, indicating a direct maternal genetic link.<br>▪ In Sweden, children often reside with their biological mothers after separation (3, 4). Therefore, mHS experience the same shared environment provided by their mother. |
| **Paternal Half-Siblings (pHS)** | 0.25 | 0 | 0 | ▪ For half-siblings, the coefficient for DG is half of the DG for full siblings because they only share one biological parent.<br>▪ MGN is set to 0 because pHS do not share the same biological mother, indicating no maternal genetic link.<br>▪ MCE is set to 0 because pHS do not experience the same shared environment provided by their mothers. |
| **Maternal Parallel Cousins (mPC)** | 0.125 | 0.5 | 0 | ▪ Cousins share one set of grandparents but not parents. Since each grandparent contributes 50% of their genes to their offspring (the parents of the cousins), and each parent passes on 50% of their genes to their child, cousins inherit approximately 12.5% of their genes from each of their shared grandparents. Therefore, the coefficient for DG, indicating the genetic correlation between cousins, is 0.125.<br>▪ For mPC (mothers are sisters), MGN is set to 0.5 because mPC share one biological grandmother (mother of the sisters), representing a partial maternal genetic link.<br>▪ For cousins, MCE is set to 0 because, on average, at the population level, especially within the Swedish population (5), shared environmental effects among cousins are generally considered to be minimal. |
| **Paternal Parallel Cousins (pPC)** | 0.125 | 0 | 0 | ▪ For pPC (fathers are sisters), MGN is set to 0 because pPC do not share any maternal genetic link. |
| **Cross Cousins (CC)** | 0.125 | 0 | 0 | ▪ For CC (mothers and fathers are brothers and sisters), MGN is set to 0 because pPC do not share any maternal genetic link |

FS: full siblings, mHS: maternal half-siblings, pHS: paternal half-siblings, mPC: maternal parallel cousins, CC: cross cousins.

To obtain estimates for GLMM, we used a binary threshold-linear mixed model in a Bayesian framework with a non-informative prior. We applied a Gibbs sampler implemented in thrgibbs1f90b (39) and generated a sample size of 150,000, with 50,000 burn-in, from the posterior distribution of the variance components. Then, we calculated the posterior mean as an estimate of the variance components. The residual variance was fixed during the calculation. We reported the results with 95% credible intervals using the Bayesian highest posterior density interval, which is analogous to two-sided 95% confidence intervals in frequentist statistics (40). For the fixed effects, we used the mean and standard deviation of the posterior to calculate the confidence interval.

We used Falconer threshold liability to study the impact of assortative mating on the estimate of the direct additive genetic effect. We compared the estimate of direct additive genetics between the model that considered assortative mating and the one that did not.

In Sweden, the National Birth Register accurately records maternal identity at childbirth, ensuring verification of the direct biological link between mother and child. The rate of misattributed paternity aligns with that of other countries, with current estimates at approximately 1% (41). Given this very low rate, we do not expect misattributed paternity to have a meaningful impact on our results.

## 3. Results

After removing multiple births (e.g., twins) and the individuals who emigrated (Figure 2), the cohort contained 982,554 individuals (48% female, Table 2). The overall ADHD prevalence across all birth cohorts in our study was 6.8% (Table 2). We observed a trend in ADHD prevalence by birth year, with an initial increase from 1992 to 1999, peaking at 7.9%, followed by a slight decline in later cohorts (Figure 3).

**Figure 2.**
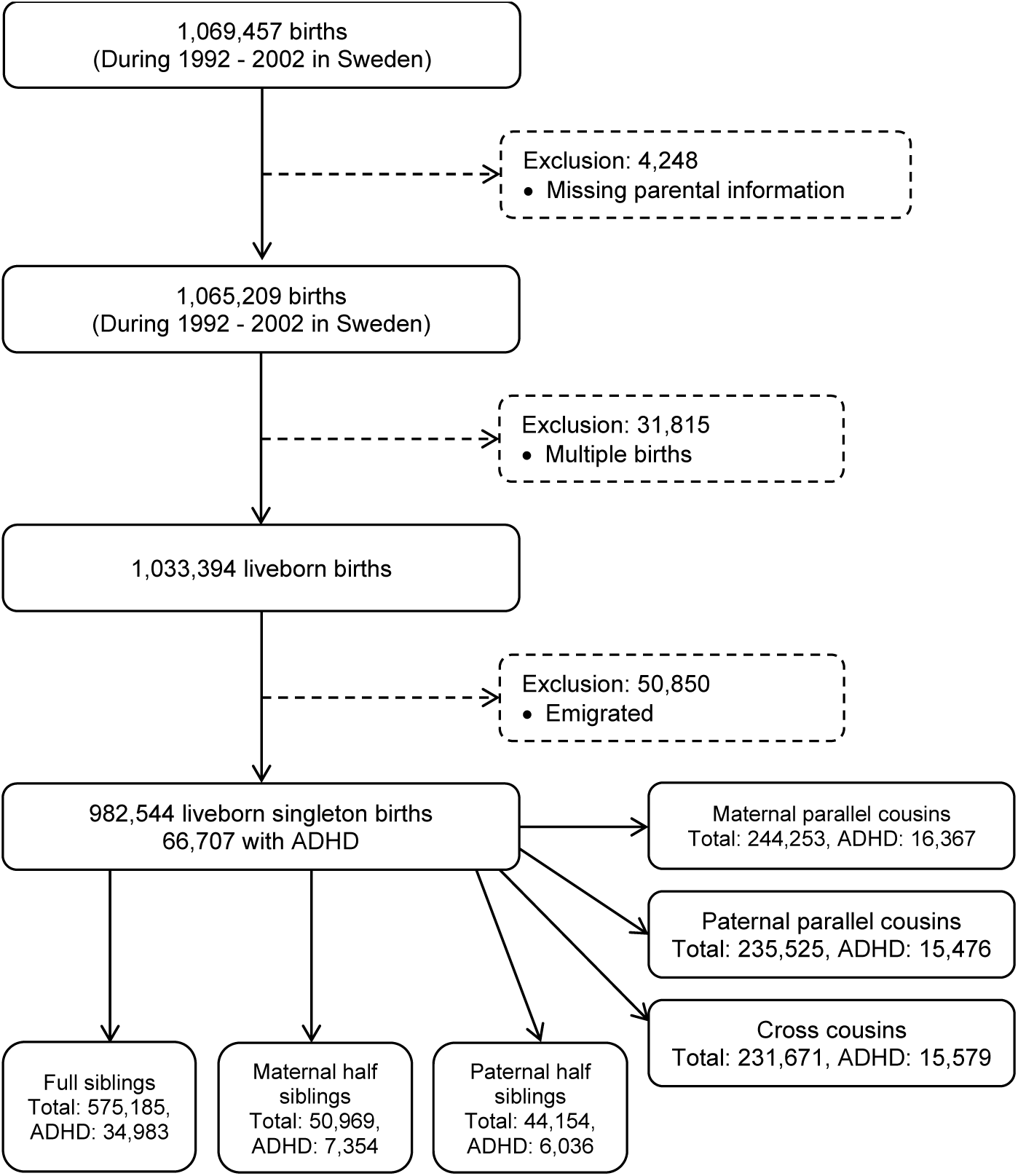
Study sample flow diagram

**Figure 3.**
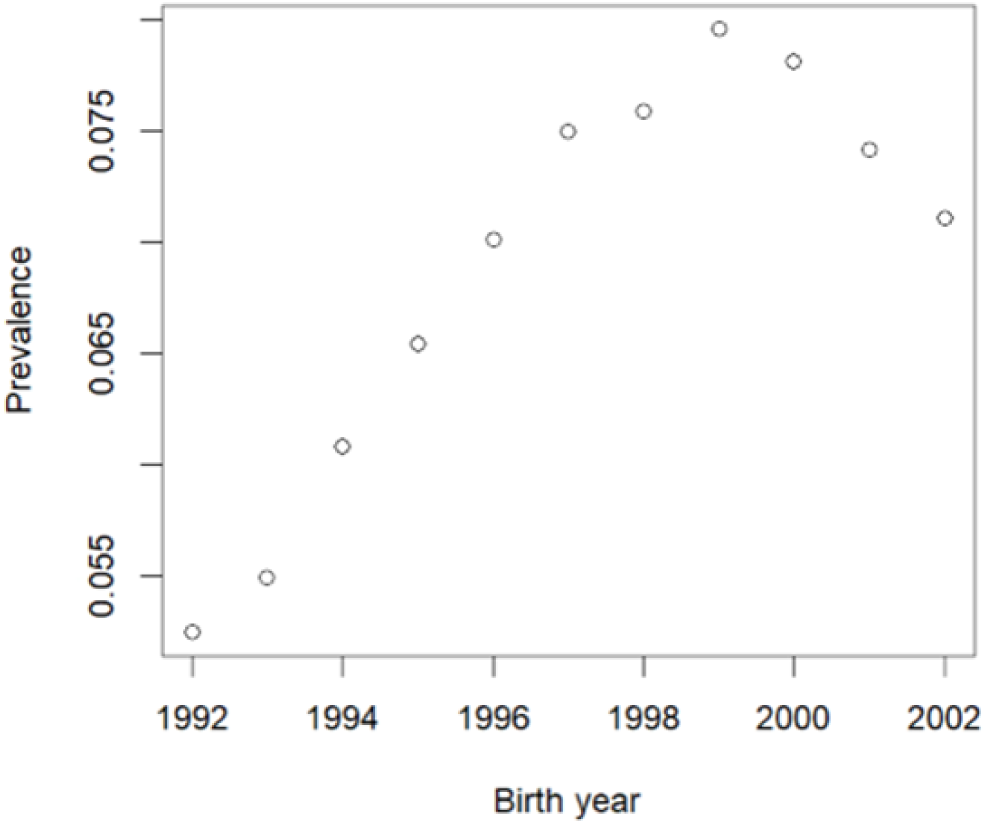
Prevalence of ADHD diagnoses by birth year for individuals born between 1992 and 2002. The prevalence increased steadily from 1992 to 1999, peaking at approximately 7.9%, followed by a decline from 2000 to 2002. The initial increase potentially reflects broader societal and diagnostic changes, such as heightened awareness and improved screening practices. The subsequent decline is attributable to right censoring, where later cohorts had less time to reach the typical diagnostic age by the end of follow-up in 2018, leading to an underestimation of prevalence in those cohorts.

**Table 2.** Study cohort including individuals born in Sweden between 1992 and 2002.

|  | Total | Siblings |  |  | Cousins |  |  |
| --- | --- | --- | --- | --- | --- | --- | --- |
|  |  | Full | mHS | pHS | mPC | pPC | CC |
| **Participants** | 982,544 | 575,185 | 50,969 | 44,154 | 244,253 | 235,425 | 231,671 |
| Male | 506,168<br>(52%) | 296,686<br>(52%) | 26,319<br>(52%) | 22,686<br>(51%) | 125,553<br>(51%) | 121,601<br>(52%) | 119,562<br>(52%) |
| Female | 476,376<br>(48%) | 278,499<br>(48%) | 24,650<br>(48%) | 21,468<br>(49%) | 118,700<br>(49%) | 113,824<br>(48%) | 112,109<br>(48%) |
| **Diagnosed with ADHD** | 66,707 | 34,983 | 7,354 | 6,036 | 16,367 | 15,476 | 15,579 |
| Male | 40,845<br>(61%) | 21,358<br>(61%) | 4,535<br>(62%) | 2,340<br>(61%) | 9,932<br>(61%) | 9,559<br>(62%) | 9,528<br>(61%) |
| Female | 25,862<br>(39%) | 13,625<br>(39%) | 2,819<br>(38%) | 3,696<br>(39%) | 6,435<br>(39%) | 5,917<br>(38%) | 6,051<br>(39%) |
| **Population Frequency of ADHD** | 0.07 | 0.06 | 0.14 | 0.14 | 0.07 | 0.07 | 0.07 |
| Male | 0.08 | 0.07 | 0.17 | 0.16 | 0.08 | 0.08 | 0.08 |
| Female | 0.05 | 0.05 | 0.11 | 0.11 | 0.05 | 0.05 | 0.05 |
| Diagnosis age | 14.7 (4.1) | 14.8 (4.0) | 14.3 (4.1) | 14.4 (4.2) | 14.7 (4.1) | 14.8 (4.1) | 14.6 (4.1) |

Abbreviations: mHS: maternal half-siblings, pHS: paternal half-siblings, mPC: maternal parallel cousins, pPC: paternal parallel cousins, CC: cross-cousins. Individuals can be included in more than one type
Note: An individual may belong to multiple family relationships, resulting in the total count across all categories exceeding the total number of individuals.

To evaluate potential bias related to outpatient data availability, we compared outpatient diagnosis frequencies between two cohorts: individuals born in 2001–2002 and those born in 1996–2000. Diagnosis frequencies were 7.5% for the later cohort and 7.2% for the earlier cohort, reflecting a slight underestimation of 0.3% for individuals born in 1996–2000.

We noted an increased population frequency in both maternal and paternal half siblings (mHS and pHS), at 0.14, twice the frequency observed in the general population (Table 2). To understand this elevation, we investigated the association between ADHD diagnosis and sibling type. Our regression analysis revealed a negative association between ADHD diagnosis and having at least one full sibling (estimate: −0.02, p < 0.001), whereas a positive association was observed with having a maternal or paternal half-sibling (estimate: 0.08 for mHS, 0.07 for pHS; both p < 0.001). This suggests that families with half-siblings who have at least one sibling with ADHD are often larger, contributing to a heightened prevalence of ADHD within those family structures.

We used Cox regression to compare the risk among different family relationship types (Table 3). The relative risk of recurrence between full siblings was 5.17 (95% CI, 4.99-5.35). Maternal half-siblings had higher relative recurrence risk as compared to paternal half-siblings (2.22 vs 1.79; Table 3). Likewise, maternal parallel cousins carried higher risk, as compared to paternal parallel cousins (2.13 vs 1.81) and cross cousins (2.13 vs 1.81). Analysis of familial risk - the probability that an individual has an affected relative of a specific type - revealed a similar pattern (Table S1).

**Table 3.** Relative Recurrence Risk (RRR) of ADHD for different relation types.

| Family Types | RRR | 95% CI |
| --- | --- | --- |
| Full siblings | 5.17 | (4.99,5.35) |
| Maternal half-siblings | 2.22 | (2.06,2.39) |
| Paternal half-siblings | 1.79 | (1.65,1.94) |
| Maternal parallel cousins | 2.13 | (2.00,2.26) |
| Paternal parallel cousins | 1.81 | (1.70,1.92) |
| Cross cousins | 1.82 | (1.74,1.91) |

We used GLMM to partition the phenotypic variation of ADHD into direct genetic effect, maternal genetic nurture effect, and maternal common envrionment effect. The model that best explained the data was direct genetic effect + maternal genetic nurture effect, as determined by Bayes Factor analyses (Table 4 and the supplementary material, section S2 and Table S5). Based on this model, 66.1% of the liability for ADHD was the result of direct additive genetic effect (95% credible interval, 0.647-0.676) and 14% from maternal genetic nurture effect (95% credible interval, 0.136-0.151). The maternal common environment effect was zero or close to zero. Using the Falconer threshold liability method to evaluate the sensitivity of the results (Supplement, section S1), we estimated 74.7% direct additive genetics and 6.7% maternal genetic nurture effect (Table S3).

**Table 4.**
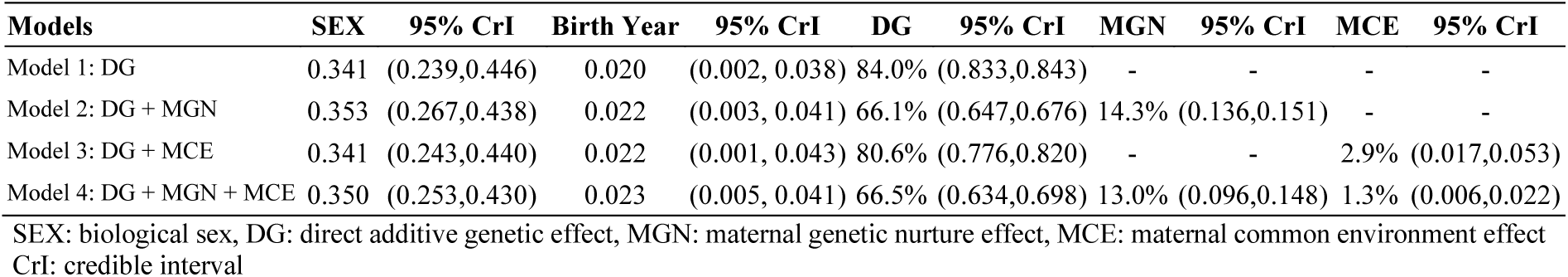
The proportion of phenotypic variance explained by different models and estimated coefficient for the fixed effect.

| Models | SEX | 95% CrI | Birth Year | 95% CrI | DG | 95% CrI | MGN | 95% CrI | MCE | 95% CrI |
| --- | --- | --- | --- | --- | --- | --- | --- | --- | --- | --- |
| Model 1: DG | 0.341 | (0.239,0.446) | 0.020 | (0.002, 0.038) | 84.0% | (0.833,0.843) | - | - | - | - |
| Model 2: DG + MGN | 0.353 | (0.267,0.438) | 0.022 | (0.003, 0.041) | 66.1% | (0.647,0.676) | 14.3% | (0.136,0.151) | - | - |
| Model 3: DG + MCE | 0.341 | (0.243,0.440) | 0.022 | (0.001, 0.043) | 80.6% | (0.776,0.820) | - | - | 2.9% | (0.017,0.053) |
| Model 4: DG + MGN + MCE | 0.350 | (0.253,0.430) | 0.023 | (0.005, 0.041) | 66.5% | (0.634,0.698) | 13.0% | (0.096,0.148) | 1.3% | (0.006,0.022) |

SEX: biological sex, DG: direct additive genetic effect, MGN: maternal genetic nurture effect, MCE: maternal common environment effect CrI: credible interval

In comparison, using a model with only direct genetics, 84% - 88% of the phenotypic variation was estimated to be the result of direct genetic transmission (Table S3).

We observed that individuals with ADHD exhibited significant assortative mating, indicating a higher likelihood of selecting partners who also have ADHD than would be expected by chance. This is quantified by the phenotypic correlation of partners (ρ, between 0 and 1), which measures the degree to which similar traits are found in partners. For a model considering only DG, we estimated ρ to be 0.3, with DG estimated at 68.0% (Table S4). This moderate positive correlation suggests that individuals with ADHD are more likely to pair with partners who also exhibit ADHD traits, rather than pairing randomly. Additionally, for a model incorporating both DG and MGN, we estimated ρ to be 0.5, indicating an even stronger phenotypic similarity among partners. This model attributed 54% of the ADHD variance to DG and 6.3% to MGN (Table S4) (for more details, see Supplementary Online Content, page 2). In both modles, we observed a significant decrease in the estimate of DG.

## 4. Discussion

This is the first study to quantify the total contribution of maternal effect, including maternal genetic nurture effect and maternal common environment effect, to the risk of ADHD. We employed a family-based quasi-experimental design to estimate these effects without the need for randomization.

As an impetus to this study, we observed that the relative recurrence risk of ADHD in maternal half-siblings was larger than in paternal half-siblings, which indicated the potential for a significant maternally derived contribution to ADHD risk architecture. To further dissect this contribution and differentiate between maternal common environment effect and maternal genetic nurture effect, we examined maternal parallel cousins. These cousins, who are genetically linked through their mothers, showed a higher risk of ADHD compared to other cousin types. This finding suggested the presence of maternal genetic nurture effect.

To accurately quantify the maternal effect, we employed a GLMM. GLMMs are well-suited for dissecting phenotypic variation into genetic and non-genetic (environmental) components due to their capability to integrate both fixed effects, which can include specific measured environmental and genetic factors, and random effects, which account for unobserved heterogeneity arising from genetic variance and unmeasured environmental influences such as maternal effect. This methodology enabled us to accurately estimate the contributions of direct genetic effect, maternal genetic nurture effect, and maternal common environment effect to the risk of ADHD. We found that direct genetic effect, or narrow-sense heritability, accounts for 67% of the variance in ADHD risk, indicating a strong genetic component. Maternal genetic nurture effect explains an additional 14% of the variance, underscoring the significant role of maternal genetics. Maternal common environment effect were found to be negligible. Narrow-sense heritability refers to the proportion of phenotypic variance attributable to the additive genetic effect, whereas broad-sense heritability encompasses the total genetic effects (additive, dominant/recessive, and epistatic).

Our findings align with those of Eilertsen et al., (42), who investigated parental contributions to externalizing problems in a broader phenotypic approach (in contrast to our study that focused on ADHD diagnosis rather than symptoms of ADHD), including symptoms of ADHD, oppositional defiant disorder, and conduct disorder at eight years of age, using genotyped data from parent-offspring trios (42). This study found that the combined indirect genetic effects from both parents (i.e., combined maternal and parental effects) accounted for 16.3% (SE = 7.9%) of the variance for inattention and 7.7% (SE = 7.9%) for hyperactivity.

Two previous studies investigated the association between maternal genetic nurture and the risk of ADHD (45, 46) by examining specific parental traits, such as behavior and health characteristics. These studies calculated parental polygenic risk scores for particular traits by combining the non-transmitted alleles of parents with effect sizes obtained from a previous GWAS and tested for associations between parental polygenic scores and the traits of the offspring. Pingault *et al.* examined several traits, including substance use, neuroticism, educational attainment, and cognitive performance, but found no strong evidence of genetic nurture (46). Similarly, Martin *et al.* used polygenic risk scores for a range of disorders, including ADHD, but their findings did not support elevated polygenic risk scores for non-transmitted parental alleles (45). Although these studies contribute significantly to our understanding, they also highlight inherent limitations. The polygenic risk score is computed by combining information from multiple genetic variants, typically single nucleotide polymorphisms, that have been linked to a trait in large-scale genetic studies. Although the polygenic risk scores are helpful in estimating an individual’s genetic risk for a particular trait, they do not account for all of the genetic factors that underlie the trait. In contrast, our study modeled maternal genetic nurture as a latent variable (unobserved). This approach does not restrict the analysis to a specific trait in the parents. Thus, our estimate of maternal genetic nurture captures the potential effects of all maternal phenotypes, which originate from maternal genetics, on the risk of ADHD in the offspring.

Assortative mating (non-random mating) has been reported among individuals with psychiatric disorders, including ADHD (30), which can limit the causal inference by increasing estimations of population prevalence and inflating the estimate of direct genetics. Our findings indicate that assortative mating among individuals with ADHD not only occurs at a significant rate but also amplifies the estimate of direct additive genetic contribution by 25%. This underscores the need for future research to carefully account for assortative mating when interpreting the genetic architecture of ADHD, as it may considerably skew our understanding of genetic predispositions.

In summary, we showed that maternal genetic nurture effect plays a significant role in risk for ADHD in offspring, and thus has an integral role in the mapping of ADHD risk architecture. While direct genetic contributions are substantial, the maternal phenotype—shaped in part by maternal genetics—can also affect the offspring’s phenotype beyond directly transmitted genetic risks: maternal genetic nurture effect. Examples of such maternal effect include prenatal exposures that could impact fetal brain development, such as elevated stress hormones or inflammation during pregnancy. Identification and appreciation of specific maternal factors, that comprise maternal effect and that can respond to intervention or even prevention may be a path for improving mental health outcomes in this population. Additional research is required to explore if specific prenatal factors associated with ADHD in offspring can be accounted for by maternal effect, i.e., whether these factors can increase the risk of ADHD in offspring beyond genetic transmission directly from the mother. This will help uncover the precise mechanism behind the maternal effect on the risk of ADHD.

Our previous studies have established a significant role for maternal genetic nurture effect in the risk of developing OCD (7.6%) (35) and Tourette syndrome (4.8%) (36). Given the comorbidity and shared etiology of these disorders with ADHD, it is crucial to examine whether maternal genetic nurture effect contributes to an increased likelihood of an individual developing these disorders concurrently. Future studies should explore the differential contributions of maternal genetic nurture effect to the phenotypes of ADHD, OCD, and Tourette syndrome in offspring. There is potential for pleiotropic effects, where maternal genes might affect multiple traits or phenotypes simultaneously through environmental pathways.

### Study Strength

The present study had several strengths. It leverages data from the Swedish National Medical Birth Register, which covers a large, genetically homogeneous population. This minimizes the potential for confounding due to population stratification. Moreover, this study is based on clinical diagnoses of ADHD by specialists, which helped to reduce the misclassification of cases and related biases.

In heritability models, the environmental contribution to phenotypic variance is often estimated without utilizing any direct measure of environmental factors. Based on these models, genetic heritability is estimated first and then subtracted from 100% to calculate the environmental contribution. Studies have shown this method can be inaccurate (47). For example, the estimates of heritability from twin studies can be inflated since the amount of variance attributable to the common environment may not be identical in monozygotic twins and dizygotic twins (47). When heritability is overestimated and thus appears large, it results in a correspondingly smaller estimate for the contribution of environmental factors to the phenotype since the total of genetic and environmental influences is assumed to equal 100%. In this study, we used a broader set of relationships (extended family design) to obtain a more accurate estimate of heritability. Extended family designs include family members beyond the nuclear family, such as cousins. Extended family designs work under the assumption that, if a phenotype is heritable, individuals who are more closely related to each other will be more similar in a specific trait, and each family member shares a genetic relatedness with all other family members, which can be estimated using pedigree information.

### Study Limitations

The present study had several limitations. Our analysis is constrained by the availability of outpatient data starting only in 2001, whereas inpatient psychiatric data has been available since 1973. For example, individuals born in 1992—the oldest in our cohort—had outpatient records available starting at age nine, rather than from birth. However, since ADHD is typically diagnosed between the ages of five and twelve, outpatient records beginning at age nine are sufficient to capture the vast majority of ADHD cases in this cohort. Consequently, while this discontinuity in outpatient data availability may lead to a slight underrepresentation of very early or less severe cases, it is unlikely to significantly impact the overall results or prevalence estimates

We observed a decline in ADHD prevalence for individuals born after 1999, which is likely influenced by right censoring. Right censoring occurs when individuals lack sufficient follow-up time to reach the typical diagnostic age for ADHD, resulting in an underestimation of prevalence in later cohorts. For instance, individuals born between 1999 and 2002 had fewer years of follow-up to receive an ADHD diagnosis by the end of the study period in 2018, contributing to the observed decline in prevalence. Despite these data limitations, the majority of ADHD diagnoses occur during childhood or adolescence, ensuring that most cases were captured even for individuals with shorter follow-up periods. To further address the potential bias introduced by data discontinuities and right censoring, we applied Cox regression and included birth year as a covariate in our GLMM analyses. These statistical methods accounted for varying follow-up times across cohorts, ensuring that our results accurately reflect true trends in ADHD prevalence and maternal effects.

While our study includes individuals diagnosed with ADHD at any age, it does not fully account for the changes in symptomatology or treatment engagement over time. Furthermore, our dataset, influenced by evolving diagnostic criteria and awareness of ADHD, may skew towards more severe manifestations of the disorder. This limitation hampers the study’s capacity to represent the entire spectrum of ADHD severity, particularly among adult populations.

The statistical modeling in this study quantifies maternal effects as a latent variable, capturing their overall influence on a child’s risk of developing ADHD. While this approach is effective for assessing the overall impact, it does not pinpoint specific sources of maternal effect or account for unshared environmental factors, such as maternal psychiatric disorders during pregnancy. Our findings suggest that shared environmental effects have a limited impact on the risk of ADHD. However, we have not tested for various unshared environmental effects, which could still play a significant role in the development of the disorder.

In our model, shared environmental effects among cousins are assumed to be zero or near zero. However, this assumption might also not always hold, depending on the phenotype and the population. If shared environmental effects among cousins were large, then the estimate of direct additive genetic effect, and maternal genetic nurture effect could be inflated, and the estimate of maternal common environment effect could be deflated. In this study, we conjecture that the shared environmental effects among cousins in Sweden, on average across a large population, are negligible. As a result, our assessments of variance components should be free from large biases.

We did not investigate paternal effects in this study; however, it is crucial to acknowledge their potential influence on the offspring’s risk of ADHD. Both maternal and paternal factors could interact in complex ways that significantly affect the condition. Therefore, our findings should not be interpreted to suggest that genetic factors influencing ADHD through maternal pathways are entirely distinct from those operating through paternal paths.

## Data Availability

Data from the Swedish national registers may be obtained from a third party and are not publicly available. Data cannot be shared publicly owing to restrictions by law. Data are available from the National Medical Registries in Sweden after approval by the Swedish Ethical Review Authority

## Author Contributions

Study concept and design: JDB, DEG, BM

Acquisition, analysis, or interpretation of data: JDB, DEG, BM

Drafting of the manuscript: JDB, LC, DEG, BM

Critical revision of the manuscript for important intellectual content: All authors.

Statistical analysis: BM

Obtained funding: JDB, DEG, BM Study supervision: JDB, DEG, BM

## Acknowledgments

This study was supported by a grant from the Beatrice and Samuel A. Seaver Foundation (DEG, JDB, BM); and the 2020 NARSAD Young Investigator Grant from the Brain & Behavior Research Foundation (BM, grant number 29355).

The sponsors of the study had no role in study design, data collection, data analysis, data interpretation, writing of the report, or in the decision to submit the paper for publication. The corresponding authors had full access to all data in the study and had final responsibility for the decision to submit for publication.

The computations were enabled by resources in project UPPMAX SENS-2018-588 provided by Uppsala University at UPPMAX.

