## Supplement for "A significant role for maternal genetic nurture in the risk architecture of attention-deficit/hyperactivity disorder"

***Supplemental Information***

### S1. Liability Threshold Model (LTM)

We used Falconer's Liability Threshold Model (LTM) to estimate direct additive genetic effect (DG), maternal genetic nurture effect (MGN), and maternal common environment effect (MCE) for attention-deficit/hyperactivity disorder (ADHD) (1). We described this approach in greater details previously (2). We first estimated familial risk (FR_ADHD_) - the probability that an ADHD proband has an affected relative of a specific type - for the different relationship types: full siblings (FS), maternal half-siblings (mHS): paternal half-siblings (pHS), maternal parallel cousins (mPC), and cross cousins (CC). For each relationship type, we estimated the regression coefficient (*b*) between familial risk in relatives of ADHD probands and population prevalence using formula 25.1.a in ((1), Chapter 25: 730-736):

$b =\frac{\left( \Phi^{-1}\left( 1-{FR}_{ADHD} \right)-\Phi^{-1}\left( 1-P \right) \right)P}{\Phi\left( \Phi^{-1}\left( 1-P \right) \right)}$,

where *P* is the population prevalence of ADHD, the prevalence of a trait among various types of relationships often mirrors the population prevalence. If this is not the case, we can use the prevalence rate of the trait specific to each relationship type to calculate the regression coefficient for each relationship type. $\Phi$ and $\Phi^{-1}$ are the standard normal distribution and its inverse, respectively (Table S1). *b* for each relationship type can be equated to its expectation based on the contribution of DG, MGN, and MCE to ADHD phenotype (Table 1, main text). Then, weighted least squares can be used to estimate the variance components. The weights are proportional to the number of probands in each relationship type (Table S1).

**Table S1.** Familial risk (FR_ADHD_) and estimated regression coefficients (*b*) for different relationship types

| **Relationship** | **n**^1^ | FR_ADHD_ |  | **B** |
| --- | --- | --- | --- | --- |
| FS | 34,983 | 0.2295 |  | 0.4079 |
| mHS | 7,354 | 0.2597 |  | 0.2649 |
| pHS | 6,036 | 0.2193 |  | 0.2002 |
| mPC | 16,371 | 0.1261 |  | 0.1825 |
| pPC | 15,481 | 0.1100 |  | 0.1449 |
| CC | 15,174^2^ | 0.1107^3^ |  | 0.1415 |

^1^Number of probands that have at least one relative of the designated type. ^2^Average number of sister-brother CC and brother-sister CC that have at least one relative of the designated type. ^3^Weighted average for sister-brother and brother-sister cross cousins.

FS: full siblings, mHS: maternal half-siblings, pHS: paternal half-siblings, mPC: maternal parallel cousins, pPC: paternal parallel cousins, CC: cross cousins.

We used different models to partition the liability of ADHD (Table S2). The estimate of MCE in Model 3 and Model 4 were negative, suggesting that these models were over-parameterized and MCE is zero or near zero. Using Model 2 (DG + MGN), we estimated 74.7% DG and 6.7% MGN.

**Table S2.** Estimation of the fraction of total variance explained by different variance components

|  | **Component** | | |
| --- | --- | --- | --- |
| **Models** | **DG** | **MGN** | **MCE** |
| Model 1: DG | 85.7% | - | - |
| Model 2: DG + MGN | 77.6% | 4.1% | - |
| Model 3: DG + MCE | 99.6% | - | -5.8% |
| Model 4: DG + MGN + MCE | 86.4% | 14.9% | -16.1% |

DG: direct additive genetic effect, MGN: maternal genetic nurture effect, MCE: maternal common environment effect.

We examined the effect of assortative mating among parents with ADHD children. Assortative mating is measured by the phenotypic correlation of partners, ρ, as shown in Table S3 (Table 7.4 in Lynch and Walsh) ^1^.

**Table S3.** Coefficients of correlation between relatives when assortative mating occurs.

| Relationship^1^ | DG | MGN | MCE |
| --- | --- | --- | --- |
| FS | 0.5(1+ρh^2^) | 1 | 1 |
| mHS | 0.25(1+2ρh^2^+ρ^2^h^2^) | 1 | 1 |
| pHS | 0.25(1+2ρh^2^+ρ^2^h^2^) | 0 | 0 |
| mPC | 0.125(1+ρh^2^)^3^ | 0.5 | 0 |
| pPC | 0.125(1+ρh^2^)^3^ | 0 | 0 |
| CC | 0.125(1+ρh^2^)^3^ | 0 | 0 |

^1^FS: full siblings, mHS: maternal half-sibings, pHS: paternal half-siblings, mPC: maternal parallel cousins, pPC: paternal parallel cousins, CC: cross cousins, ρ: coefficient of assortative mating.

Because the influence of assortative mating is based on both the heritability estimate, h^2^, and the assortative mating parameter, ρ, to obtain estimates for the variance components as well as the assortative mating parameter ρ, the system of equations needs to be solved iteratively. We performed a grid search for ρ and chose the parameter that resulted in the highest adjusted R^2^ for model fit. The best fit for Model 1 was for ρ = 0.3 with DG = 68.0% (Table S4). The estimate for DG was lower (62.3%) when taking assortative mating into consideration, as compared to the model that did not consider it, where the estimate was 87.9%. These results are expected since similarities between relatives are inflated in the presence of assortative mating. The best fit for Model 2 was for ρ = 0.5 with DG = 54% and MGN = 6.3% (Table S4). Similarly, in Model 2, we observed a significant decrease in the DG estimate, while the MGN estimate remained relatively stable.

**Table S4.** Estimation of the fraction of total variance explained by different variance components for different assortative mating rates.

|  |  | **Model 1: DG** | **R^2^** | **Model 2: DG + MGN** | | **R^2^** |
| --- | --- | --- | --- | --- | --- | --- |
| ρ |  | **DG** |  | **DG** | **MGN** |  |
| 0 |  | 85.7% | 0.974 | 77.6% | 4.1% | 0.960 |
| 0.1 |  | 78.4% | 0.986 | 72.3% | 3.6% | 0.983 |
| 0.2 |  | 72.7% | 0.992 | 67.1% | 3.8% | 0.991 |
| **0.3** |  | **68.0%** | **0.995** | 62.1% | 4.5% | 0.995 |
| 0.4 |  | 64.0% | 0.994 | 57.8% | 5.4% | 0.997 |
| **0.5** |  | 63.0% | 0.991 | **54.0%** | **6.3%** | **0.998** |
| 0.6 |  | 57.6% | 0.988 | 50.6% | 7.2% | 0.997 |
| 0.7 |  | 54.9% | 0.983 | 47.7% | 8.0% | 0.996 |
| 0.8 |  | 52.4% | 0.977 | 45.1% | 8.9% | 0.994 |
| 0.9 |  | 50.3% | 0.970 | 42.9% | 9.7% | 0.992 |
| 1 |  | 48.4% | 0.963 | 41.0% | 10.4% | 0.991 |

DG: direct additive genetic effect, MGN: maternal genetic nurture effect, MCE: maternal common environment effect, ρ: coefficient of assortative mating.

### S2. Fitting variance components model

We used Bayesian binary threshold-linear mixed models with a non-informative prior. For each model, we obtained an estimate for log marginal likelihood. To compare two models, H_1_ and H_0,_ we calculated Log Bayes Factor (BF), where

Log(BF) = Log(marginal density for Bayes factor for H_1_) - Log(marginal density for Bayes factor for H_0_).

If Log(BF) is larger than 1, then H_1_ fits the data better than H_0_. The results for different models are illustrated in Table S5. We reported the results with 95% credible intervals (CrI) using the Bayesian highest posterior density interval, which is analogous to two-sided 95% CIs in frequentist statistics.

**Table S5.** Estimates of the fixed effect and the variance components based on different models

| **Models** | **SEX** | **95% CrI** | **Birth Year** | **95% CrI** | **DG** | **95% CrI** | **MGN** | **95% CrI** | **MCE** | **95% CrI** | **Log(MD)** |
| --- | --- | --- | --- | --- | --- | --- | --- | --- | --- | --- | --- |
| Model 1: DG | 0.341 | (0.239,0.446) | 0.020 | (0.002, 0.038) | 84.0% | (0.833,0.843) | - | - | - | - | -1396454 |
| Model 2: DG + MGN | 0.353 | (0.267,0.438) | 0.022 | (0.003, 0.041) | 66.1% | (0.647,0.676) | 14.3% | (0.136,0.151) | - | - | -1396352 |
| Model 3: DG + MCE | 0.341 | (0.243,0.440) | 0.022 | (0.001, 0.043) | 80.6% | (0.776,0.820) | - | - | 2.9% | (0.017,0.053) | -1396525 |
| Model 4: DG + MGN + MCE | 0.350 | (0.253,0.430) | 0.023 | (0.005, 0.041) | 66.5% | (0.634,0.698) | 13.0% | (0.096,0.148) | 1.3% | (0.006,0.022) | -1396710 |

SEX: biological sex, DG: direct additive genetic effect, MGN: maternal genetic nurture effect, MCE: maternal common environment effect

Log(MD): Log(marginal density for Bayes factor), CrI: credible interval, MD: Marginal density
